# COVID-19 Wastewater Epidemiology: A Model to Estimate Infected Populations

**DOI:** 10.1101/2020.11.05.20226738

**Authors:** Christopher Steven McMahan, Stella Self, Lior Rennert, Corey Kalbaugh, David Kriebel, Duane Graves, Jessica A. Deaver, Sudeep Popat, Tanju Karanfil, David L. Freedman

## Abstract

**BACKGROUND:** Wastewater-based epidemiology (WBE) provides an opportunity for near real-time, cost-effective monitoring of community level transmission of SARS-CoV-2, the virus that causes COVID-19. Detection of SARS-CoV-2 RNA in wastewater can identify the presence of COVID-19 in the community, but methods are lacking for estimating the numbers of infected individuals based on wastewater RNA concentrations.

**METHODS:** Composite wastewater samples were collected from three sewersheds and tested for SARS-CoV-2 RNA. A Susceptible-Exposed-Infectious-Removed (SEIR) model based on mass rate of SARS-CoV-2 RNA in the wastewater was developed to predict the number of infected individuals. Predictions were compared to confirmed cases identified by the South Carolina Department of Health and Environmental Control for the same time period and geographic area.

**RESULTS:** Model predictions for the relationship between mass rate of virus release to the sewersheds and numbers of infected individuals were validated based on estimated prevalence from individual testing. A simplified equation to estimate the number of infected individuals fell within the 95% confidence limits of the model. The unreported rate for COVID-19 estimated by the model was approximately 12 times that of confirmed cases. This aligned well with an independent estimate for the state of South Carolina.

**CONCLUSIONS:** The SEIR model provides a robust method to estimate the total number of infected individuals in a sewershed based on the mass rate of RNA copies released per day. This overcomes some of the limitations associated with individual testing campaigns and thereby provides an additional tool that can be used to better inform policy decisions.

## INTRODUCTION

Early detection and containment of the novel severe acute respiratory syndrome coronavirus 2 (SARS-CoV-2) is essential to containing community outbreaks of the Coronavirus Disease 2019 (COVID-19).^1^ Clinical testing of every individual in a community is impractical and expensive.^2^ By contrast, environmental surveillance may enable continuous, cost-effective means of monitoring communities for early warning and the progress of outbreaks. Wastewater-based epidemiology (WBE) is a promising tool to assess COVID-19.^3^ A wide range of WBE applications exist, including detection of illicit drugs, surveillance of pathogenic enteric viruses, and assessment of potential industrial chemical exposures.^4^ Both symptomatic and asymptotic individuals shed SARS-CoV-2 in their feces, and studies have demonstrated detection of SARS-CoV-2 genes in raw sewage and primary sewage treatment sludge.^5–12^

Before WBE can be widely adapted for COVID-19 prevention and management, a method is needed to estimate the number of active infections from the viral RNA load detected in wastewater. To capture the infection dynamics of COVID-19, we employed a Susceptible-Exposed-Infectious-Recovered (SEIR) model, which has been previously used to predict SARS-CoV-2 transmission.^13–15^ The SEIR model utilized here estimated the number of infections based on the mass rate of virus RNA in sewage (i.e., gene copies d^-1^) for two of the three wastewater treatment plants (WWTPs) surveilled, accounting for variability in factors such as fecal production rate, SARS-CoV-2 RNA density in feces, and decay rates during transit in sewer lines. Expressing viral RNA load in terms of a mass rate minimizes the impact of dilution of sewage from non-viral sources such as stormwater runoff and infiltration during rain events. This provides a platform for comparing results across sewersheds in any location.

## METHODS

### SEWERSHEDS

Wastewater from three sewersheds was monitored (Fig. 1): The Clemson University (CU) wastewater treatment plant (WWTP) serves the main campus with a student body of ∼25,000 and a small adjacent residential subdivision. The Cochran Road WWTP serves approximately one-half of the City of Clemson, including a high density of off-campus student housing. The Pendleton/Clemson WWTP serves the Town of Pendleton and the other half of the City of Clemson and consists of a mixture of residential neighborhoods and off-campus student housing. The Cochran Road and Pendleton/Clemson WWTP sewer sheds align closely with the 29631 zip code, for which the South Carolina Department of Health and Environmental Control (SCDHEC) reports COVID-19 cases (https://scdhec.gov/covid19/sc-testing-data-projections-covid-19). There are no major industrial dischargers.

**Figure 1.**
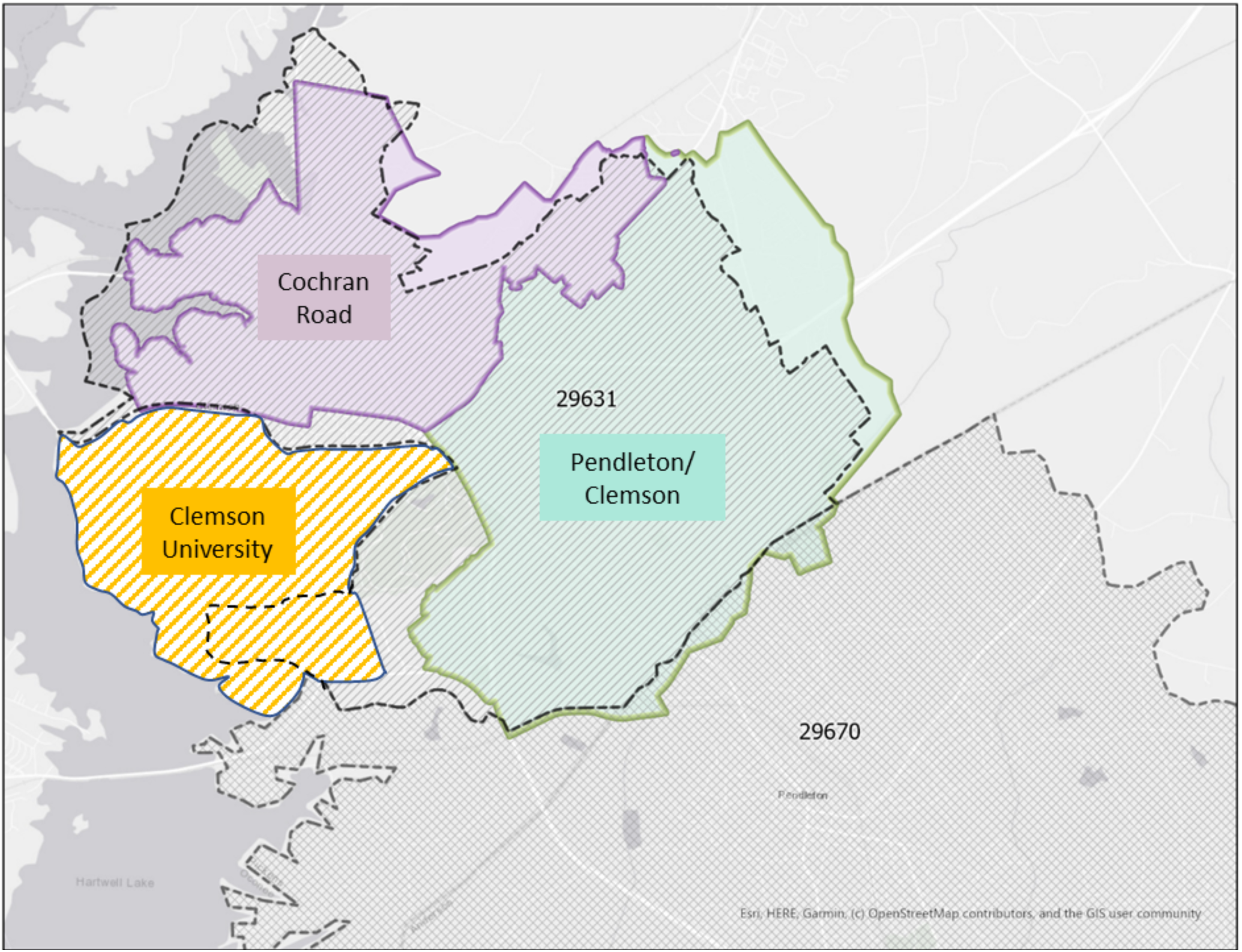
Sewersheds under surveillance for SARS-CoV-2 in wastewater. The 29631 zip code (dashed black line) overlaps mainly with the Cochran Road and Pendleton/Clemson sewersheds. The Clemson University sewershed encompasses the campus and a small residential subdivision adjacent to the campus.

### SAMPLE COLLECTION, SARS-CoV-2 DETECTION AND QUANTIFICATION

Sewage samples were collected weekly or bi-weekly starting at the end of May from the CU WWTP and mid-June from Cochran Rd and Pendleton/Clemson. Composite samples were collected continuously over a 24-hour period and stored at ∼4 °C. Samples (500 mL in Nalgene bottles) were shipped on ice overnight to SiREM Laboratory (Knoxville, TN) for quantification of SARS-CoV-2. Details on the methods used to quantify SARS-CoV-2 in wastewater samples are provided in the Supplementary Appendix. Quantification of gene copies/L was performed using the N protein gene. Detection limits ranged from 860 to 4,000 copies L^-1^.

### MODELING INFECTED INDIVIDUALS

The SEIR model organizes individuals into four compartments according to their disease status and other criteria: susceptible (individuals who may acquire the infection); exposed (infected individuals who are not yet contagious); infectious (individuals who are infected and contagious); and recovered (those who are no longer infectious).^16^ Recovered individuals are assumed to be no longer susceptible, although this assumption can be easily modified if future evidence contradicts this. Let *S*(*t*), *E*(*t*), *I*(*t*), and *R*(*t*) denote the proportion of the population that is susceptible, exposed, infectious, and recovered, respectively, at time *j*. The transitions between compartments are governed by the following system of differential equations:

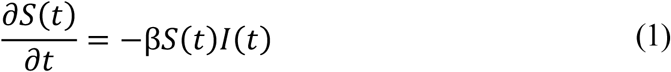

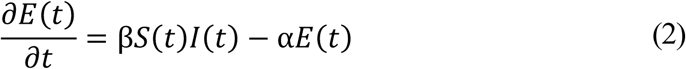

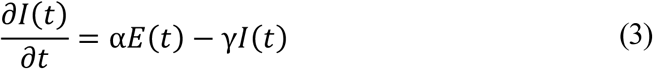

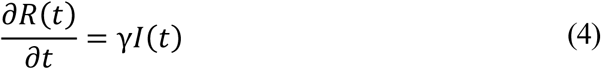

where *β* is the number of contacts per day that are sufficient to lead to infection, *α* is the rate of progression from exposed to infectious (the reciprocal is the incubation period), and *γ* denotes the rate of loss of infectiousness (the reciprocal is the infectious period). As is common, and without loss of generality, we assume that time is measured in days and all associated rate parameters are interpreted accordingly. In particular, the basic reproduction number, *R*_0_, for an SEIR model is given by *R*_0_ = *βS*(0)*/γ*, where *S*(0) denotes the proportion of the population that is initially susceptible.^17^ Hethcote^16^ and Abou-Ismail^18^ provide an overview of SEIR models and related variants.

For this study, an α value of 1/5 was assumed, as the median incubation period of COVID-19 is 5 days.^19^ The recovered rate (γ) was set to 1/10, since a typical infectious period for COVID-19 is 10 days.^20^ We assumed that 0.005% of the population was initially infected, another 0.005% of the population was initially exposed, and the remaining 99.99% of the population was initially susceptible. Three values of β were evaluated: 0.15, 0.20 and 0.25, corresponding to R_0_ values of 1.50, 2.0, and 2.5, respectively, consistent with reported estimates for COVID-19.^21–23^

Using the SEIR model under the aforementioned parameter settings, we assumed that the number of new infections on the *j*^*th*^ day (denoted *C*_*j*_) obeys:

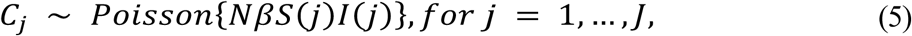

where *N* is the number of individuals in the sewershed. For each day *j* = 1, …, *J*, we sampled *C*_*j*_ from the distribution specified above. For *i* = 1, …, *C*_*j*_, let *V*_*ij*_(*t*) denote the number of copies of SARS-CoV-2 RNA entering the sewershed through the feces of the *i*^*th*^ individual from among the *C*_*j*_ who became infected on day *j*. We refer to *V*_*ij*_(*t*) as the ‘viral trajectory’ of individual (*i, j*). Specifically, we assumed

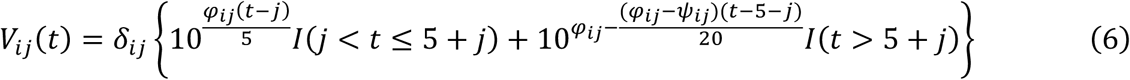

where *δ*_*ij*_ is the number of grams of feces contributed by *i*^th^ individual who was infected on the *j*^th^ day, *φ*_*ij*_ is the log_10_ maximum RNA copies per gram of feces being shed (assumed to occur 5 days after being infected), and *ψ*_*ij*_ is the log_10_ RNA copies per gram of feces being shed 25 days after being infected. This model of viral trajectory assumes an incubation time of 5 days; i.e., the number of days until symptoms appear.^19^ During this period, the individual’s viral shedding is allowed to increase to its maximum. The maximum was determined based on findings of Wölfel et al.^24^ and the evidence that maximum shedding of SARS-CoV-2 occurs around the onset of symptoms.^25^ To control the decline of viral shedding over the course of the infection, we further assimilated findings from Wölfel et al.^24^ to set the log_10_ RNA copies per gram of feces at 25 days post infection (20 days post symptoms). The specific settings are *log*_,10_(δ_*ij*_) ∼ *N*(2.11,0.25^2^), φ_*ij*_ ∼ *N*(7.6,0.8^2^), and ψ_*ij*_ ∼ *N*(3.5,0.35^2^). Given equation (6), the viral load being introduced into the sewershed at time *t* is given by:

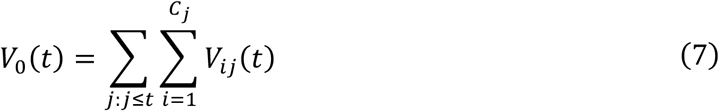

To relate the viral load being introduced into the system to that being measured, we account for time dependent degradation in the sewer system using the exponential decay model discussed by Hart and Halden.^2^ Accordingly, the measured viral load at time *t* is given by:

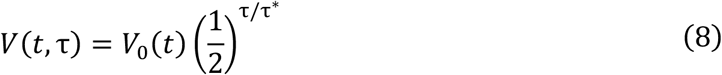

where *τ* denotes the holding time (i.e., elapsed time between waste excretion and arrival at the wastewater treatment plant), *V* (*t, τ*) is the ‘downstream’ RNA copies measured at the wastewater treatment plant, *V*_0_(*t*) is the viral load introduced into the sewershed, and *τ*\* is the temperature dependent half-life. Based on sewage residence times cited in Kapo et al.^26^ for systems with capacities of ∼3.8E6 L d^-1^, *τ* for both sewersheds in this study was set at 1.1 h. The temperature adjusted half-life is determined by:

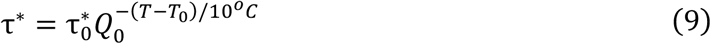

where *τ* _0_ * is the half-life (hr) at an ambient temperature of *T*_0_, *T* is the current temperature of the system, and *Q*_0_ is a temperature-dependent rate of change.^27^ Ahmed et al.^28^ report that for SARS-CoV-2, *τ* _0_ *∈{57, 202}at 20^*o*^*C*, and that *Q*_10_ ∈ {2,3}. Based on these values, we sampled *τ* _0_ * ∼*N* (130, 25^2^) and *Q*_0_ ∼*N* (2.5, 0.15^2^) during Monte Carlo simulations.

Code that implements the full model was curated into an R package, available via GitHub at (https://github.com/scwatson812/COVID19WastewaterModel). This code can be used to conduct the full Monte Carlo study described herein. A simplified version of the model was packaged as an R Shiny application (https://rennertl.shinyapps.io/Wastewater_projections/). This application can provide real time assessments in a user-friendly fashion. As a part of both of these sets of code, users can specify input parameters (e.g., reproductive number, viral half-life, sewage temperature, etc.) and re-run analyses adaptable to any sewershed.

A simplified estimate of infected individuals on any given day, based on the mass rate of virus copies present in sewage (numerator) and the mass rate for shedding of the virus (denominator) is:

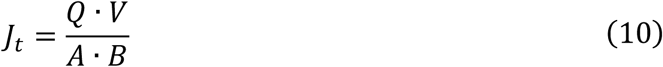

where *J*_*t*_ is the number of infected individuals for a given 24 h period, *Q* is the average flow rate at the WWTP (L d^-1^) for a 24 h period, *V* is the virus copies per liter, *A* is the rate of feces production per person (g d^-1^), and *B* is the maximum rate at which the virus is shed (RNA copies g feces^-1^ d^-1^). *Q* was obtained from records kept by the WWTPs; *V* was quantified in samples sent to SiREM (RNA copies L^-1^); *A* was set at 128 g d^-1^ the median value for developed countries^29^; and *B* was set at 4.7E07 RNA copies per g feces, the maximum rate reported by Wölfel et al.^24^ The RNA mass rate is the numerator in equation 10, i.e., a product of the sewage flow rate and virus RNA concentration.

## RESULTS

### SARS-CoV-2 SURVEILLANCE DATA

In anticipation of students returning to campus for the Fall 2020 semester, CU began surveillance for SARS-CoV-2 at its WWTP in late May (Table 1). At the time, only essential staff and administrators were using the campus, along with a limited number of graduate students and student athletes; no other undergraduates were present. Consequently, the flow rate through the treatment plant was ∼one-third to one-half of what typically occurs when campus access is unrestricted. The intent of monitoring during this period was to establish a baseline for comparison to operation when undergraduate students returned in mid-September. For the duration of the surveillance reported here, virus copies were close to or below detection.

**Table 1.** SARS-CoV-2 RNA levels in three adjoining sewersheds.

| Date<sup>a</sup> | Rainfall (cm) | CU WWTP |  |  | Cochran Road WWTP |  |  | Pendleton/Clemson WWTP |  |  | Zip Code 29631 |  |  |
| --- | --- | --- | --- | --- | --- | --- | --- | --- | --- | --- | --- | --- | --- |
|  |  | Flow Rate (10<sup>6</sup> L d<sup>-1</sup>) | RNA (copies L<sup>-1</sup>) | RNA Rate (10<sup>12</sup> copies d<sup>-1</sup>) | Flow Rate (10<sup>6</sup> L d<sup>-1</sup>) | RNA (copies L<sup>-1</sup>) | RNA Rate (10<sup>12</sup> copies d<sup>-1</sup>) | Flow Rate (10<sup>6</sup> L d<sup>-1</sup>) | RNA (copies L<sup>-1</sup>) | RNA Rate (10<sup>12</sup> copies d<sup>-1</sup>) | Flow Rate (10<sup>6</sup> L/d) | RNA Rate (10<sup>12</sup> copies d<sup>-1</sup>) | ~Infected Individuals<sup>d</sup> |
| 6/11/20 | 0.00 | 1.34 | BLD<sup>b</sup> | - | NS<sup>c</sup> | NS | NS | NS | NS | NS | - | - | - |
| 6/16/20 | 0.00 | 1.64 | 5.5E+03 | 0.0090 | 2.68 | 1.8E+06 | 4.82 | NS | NS | NS | - | - | - |
| 6/18/20 | 1.27 | 1.39 | BLD | - | 3.67 | 5.5E+05 | 2.02 | NS | NS | NS | - | - | - |
| 6/23/20 | 1.80 | 1.39 | BLD | - | 3.91 | 3.8E+06 | 14.86 | 4.16 | 2.9E+05 | 1.21 | 8.07 | 16.1 | 2,649 |
| 6/25/20 | 0.20 | 1.50 | BLD | - | 3.84 | 9.7E+05 | 3.72 | 4.23 | 9.0E+05 | 3.81 | 8.07 | 7.53 | 1,242 |
| 6/30/20 | 0.00 | 1.33 | BLD | - | 3.34 | 9.8E+05 | 3.28 | NS | NS | NS | - | - | - |
| 7/2/20 | 0.00 | 1.36 | 1.8E+04 | 0.0244 | NS | NS | NS | 3.69 | 2.9E+05 | 1.07 | - | - | - |
| 7/7/20 | 3.40 | 1.66 | 1.0E+04 | 0.0166 | 6.28 | 1.7E+05 | 1.07 | 4.02 | 2.2E+05 | 0.88 | 10.29 | 1.95 | 322 |
| 7/9/20 | 0.00 | 1.44 | BLD | - | 3.89 | 2.4E+05 | 0.93 | 3.66 | 3.3E+05 | 1.21 | 7.55 | 2.14 | 353 |
| 7/14/20 | 0.00 | 1.44 | BLD | - | 3.41 | 1.9E+06 | 6.48 | 3.48 | 4.5E+05 | 1.56 | 6.89 | 8.05 | 1,327 |
| 7/16/20 | 0.00 | 1.45 | BLD | - | 3.71 | 4.9E+05 | 1.82 | 3.47 | 1.2E+05 | 0.42 | 7.17 | 2.23 | 368 |
| 7/21/20 | 0.00 | 1.34 | 1.2E+04 | 0.0160 | 3.35 | 5.9E+04 | 0.20 | 3.45 | 2.0E+05 | 0.69 | 6.81 | 0.89 | 147 |
| 7/28/20 | 0.00 | 1.41 | 1.4E+04 | 0.0197 | 3.33 | 9.1E+05 | 3.03 | 3.39 | 1.3E+05 | 0.44 | 6.72 | 3.47 | 573 |
| 8/5/20 | 0.00 | 1.53 | BLD | - | 2.59 | 7.0E+05 | 1.81 | 3.69 | 1.6E+05 | 0.59 | 6.28 | 2.41 | 397 |
| 8/11/20 | 0.00 | 1.46 | BLD | - | 2.80 | 7.6E+04 | 0.21 | 3.89 | 1.1E+05 | 0.43 | 6.69 | 0.64 | 106 |
| 8/18/20 | 0.03 | 1.55 | 1.7E+04 | 0.0264 | 3.52 | 7.0E+04 | 0.25 | 4.17 | 1.5E+05 | 0.63 | 7.69 | 0.87 | 144 |
| 8/25/20 | 0.18 | 1.94 | BLD | - | 3.50 | 8.0E+05 | 2.80 | 4.05 | 2.3E+05 | 0.93 | 7.55 | 3.73 | 616 |

<sup>a</sup>The CU WWTP was also sampled on 5/27/20, 5/28/20, 6/2/20, 6/4/20, and 6/9/20; all results were BLD.
<sup>b</sup>BLD = below detection level.
<sup>c</sup>NS = no sample taken.
<sup>d</sup>Calculated using equation 10.

Because most CU students live off-campus, the surveillance plan included monitoring wastewater in the two WWTPs that adjoin the campus. Viral RNA levels were mostly above 10^5^ copies L^-1^ in both sewersheds. A modest decrease in concentrations occurred at the Cochran Road WWTP starting in mid-July, possibly related to passage of a mask ordinance in the City of Clemson on June 25, 2020. Table 1 also reports mass rates for RNA copies, based on flow rate (*Q*) times concentration (*V*).

### SEIR MODEL

The SEIR model (Fig. 2A) predicts decreases in the susceptible population as individuals become exposed and infected, then recover. The distribution of RNA copies per day present in sewage was estimated for the combined Cochran Road + Pendleton/Clemson sewersheds (Fig. 1), with an estimated population of 16,000 individuals (Fig. 2B). As the number of infected individuals increases, so does the mass rate of viral RNA production appearing in the sewage; as individuals recover and shedding rates decrease, the mass rate of viral copies discharged decreases.

**Figure 2.**
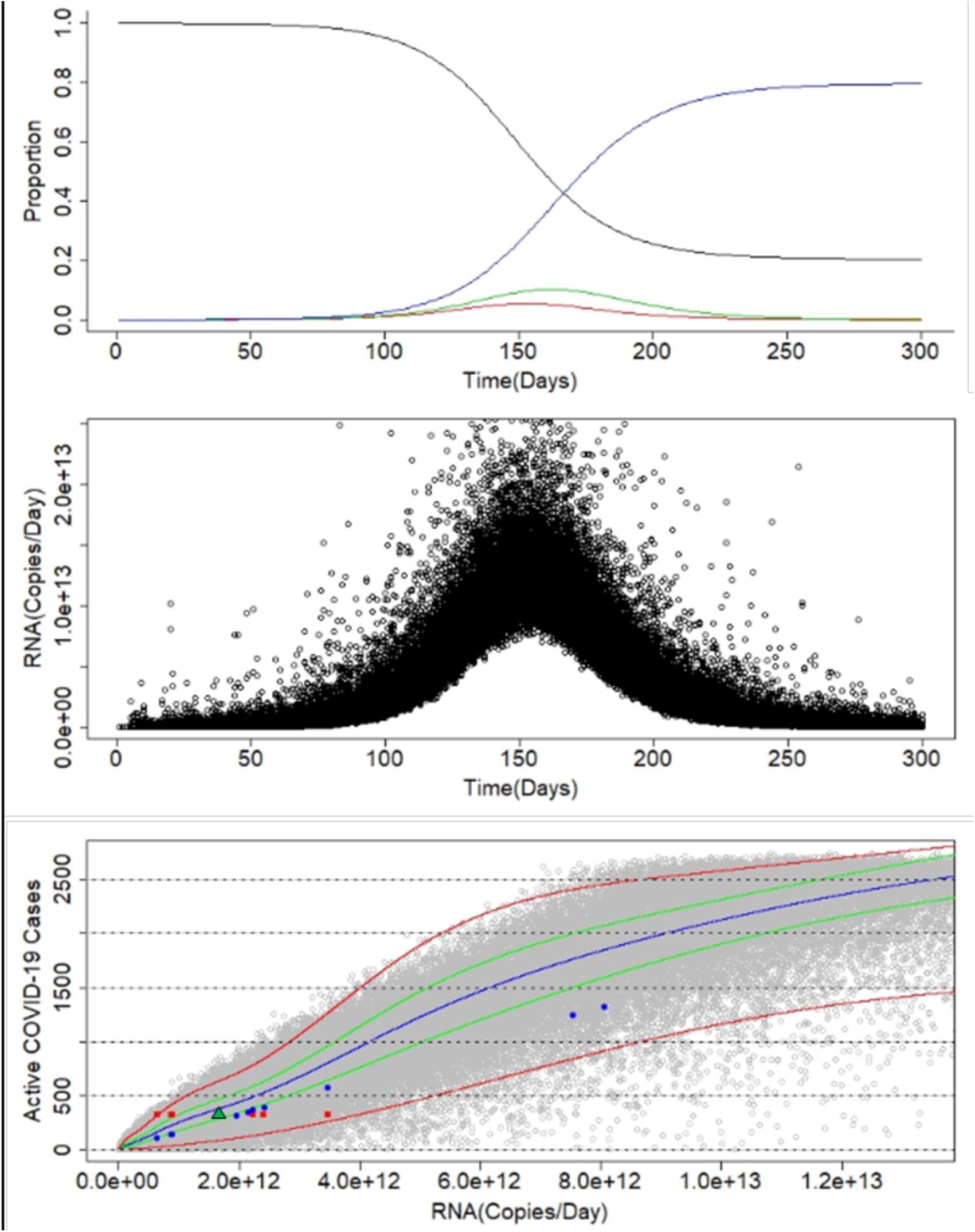
SEIR model. Panel A: Proportions of the population that are susceptible (**black**), exposed (**red**), infectious (**green**), and recovered (**blue**). Panel B: Model predictions for mass rate of SARS-CoV-2 RNA in wastewater over time. Individual black points represent each simulation. Panel C: Predictions of the number of infections versus RNA mass rate. Individual gray points represent each simulation. The **blue** line represents the median; the **green** and **red** lines represent the 75% and 95% confidence intervals, respectively. Colored data points correspond to measured RNA mass rates (Table 1) and estimates of infected individuals based on equation 10 (**blue**) and estimated positive cases (320) assuming 2% of the population was infected (**red**). The **green** triangle represents the average RNA mass rates for July 16 to August 18 (Table 1) versus the ∼320 positive cases.

Model predictions for the relationship between mass rate of virus release and numbers of infected individuals are shown in Figure 2C. Using mass rates (Table 1) for the 29631 zip code, estimates of the number of infected individuals using equation 10 are plotted (blue circles) and fall within the 95% confidence range. Equation 10 underestimated the number of infected individuals, likely because it does not take into account virus decay during transit. Another estimate of the number of infected individuals is also plotted against the measured mass rates. For a population of 16,000 and a 2% level of infection (based on individual testing of CU employees between July 20-22, 2020, many of whom live in the 29631 zip code area), the estimated number of active cases is 320. The red points in Figure 2C represent this estimate versus virus mass rates between July 16 and August 18, 2020 (Table 1); all fall within the 95% Confidence range. When these measurements are averaged, the green triangle in Figure 2C falls within the 75% confidence interval for the SEIR model.

The value of β used in the SEIR model was 0.20. β impacted the timing and magnitude of the peak of the epidemic (Fig. 2A) but did not appreciably impact the relationship between active cases and mass rate of gene copies detected in wastewater (Fig. 2C; Supplementary Appendix 1).

Model predictions based on RNA mass rates were compared to confirmed cases for the 29631 zip code (https://scdhec.gov/covid19/sc-testing-data-projections-covid-19) corrected for underreporting, based on an estimate for South Carolina from Wu et al.^30^ (15 infections for every confirmed case). SCDHEC active cases were calculated as the sum of the confirmed cases within a 15-day window formed by the previous 10 days, the current day, and the upcoming 4 days. This period was defined to match the exposed and infectious periods discussed above as well as account for the likelihood that people would seek testing at the onset of symptoms, i.e., around day 5 of being infected. The regression line for the model prediction of active cases is not statistically different (*p*<0.05) from the trend line for a perfect match between the model and cases corrected for underreporting (Fig. 3).

**Figure 3.**
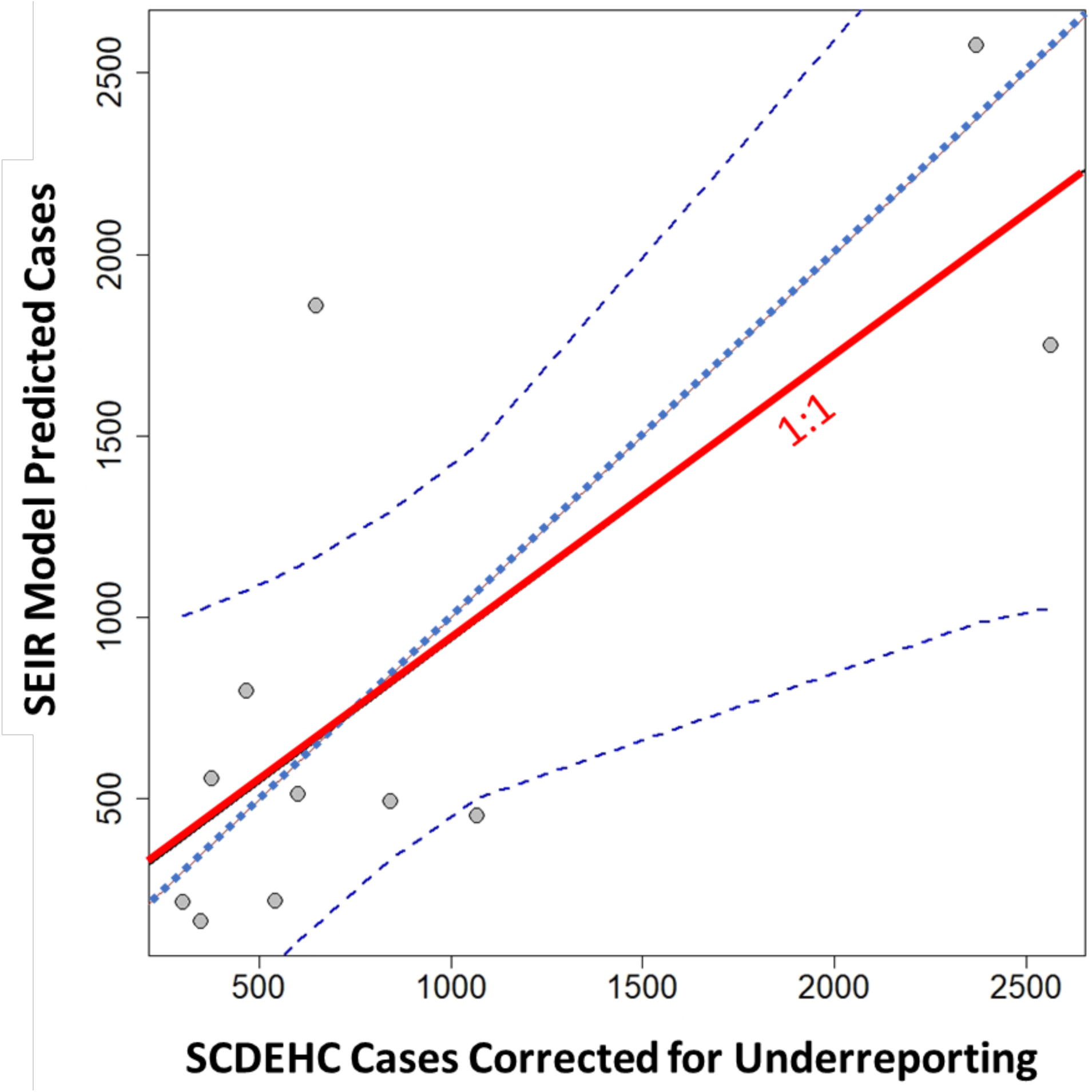
SEIR model predictions of active cases in the 29631 zip code based on RNA mass rates is wastewater versus SCDEHC cases corrected for underreporting based on data from Wu et al.^30^ The dashed blue line represents the linear regression, bounded by the 95% confidence band. The red line represents a perfect match between the model and active cases.

## DISCUSSION

Hundreds of communities worldwide are now using wastewater surveillance of SAR-CoV-2. Wastewater surveillance fills a gap left by incomplete individual testing. Nevertheless, the usefulness of WBE has been limited by the difficulty of relating the prevalence of the virus in wastewater to the number of infected individuals. The model presented here offers a method to estimate infected individuals based on the mass rate of RNA in wastewater. Mass rate is a preferred method in comparison to concentration. Sewerage collection systems in most areas are subject to dilution through stormwater runoff and infiltration, thereby lowering the concentration of virus during rain events. Use of a mass rate mitigates this impact by focusing on the product of flow rate and concentration, i.e., mass rates are unaffected by a decrease in concentration when multiplied by an increase in flow rate, and vice versa.

The utility of the SEIR model was demonstrated for two sewersheds that comprise the community adjacent to CU. Because these sewersheds coincide with the 29631 zip code, it was possible to relate model estimates of infected individuals to new cases. The correlation suggests that new cases are significantly underreported. Wu et al.^30^ estimated 15 infections for every confirmed case in South Carolina; the SEIR model predicted a ratio of 12 (95% confidence interval of 5.7–18.6). One of the contributing factors for college communities relates to students using their parent’s address as their permanent one, and most parents live outside campus communities. This further emphasizes the value of using the model to estimate infected individuals.

One of the challenges with WBE is how to communicate results to policy makers and the public. Concentration levels are difficult to comprehend and may be subject to considerable dilution in collection systems that combine sanitary waste with stormwater runoff. We propose a system that relates the percentage of people in a sewershed who are infected to the RNA copies present in sewage per person per day (Table 2). For any given sewershed, RNA copies per person per day can be calculated using the sewage flow rate times the virus RNA concentration (i.e., the RNA mass rate), divided by the number of people present. Ranges can be adjusted to sewershed-specific conditions. In the absence of widespread and systematic human-based testing, concern levels from WBE provide a way to communicate the severity of transmission to the public. For example, a concern level of II can serve as a leading indicator that active transmission is underway, even before new cases are reported.

**Table 2.** Proposed system for interpreting SARS-CoV-2 RNA levels in wastewater.

| % Infected | Copies Person<sup>-1</sup> d<sup>-1</sup> <sup>a</sup> | Concern Level |
| --- | --- | --- |
| <0.01 | <6.0E+05 | 0 |
| 0.01-0.10 | 6.0E+05 – 6.0E+06 | I |
| 0.10-1.0 | 6.0E+06 – 6.0E+07 | II |
| 1.0 – 5.0 | 6.0E+07 – 3.0E+08 | III |
| >5.0 | >3.0E+08 | IV |

<sup>a</sup>Estimated based on the percent infected times the denominator in equation 10 (i.e., $A \times B$ ).

Because infected individuals continue to shed virus after they have recovered, SARS-CoV-2 RNA levels in wastewater becomes a lagging indicator. Once levels decline sufficiently, WBE can once again be used as a leading indicator of a resurgence of transmission.

## Data Availability

All data used is publicly available and can be found in the links below.

https://scdhec.gov/covid19/sc-testing-data-projections-covid-19

https://www.clemson.edu/covid-19/testing/wastewater-dashboard.html

## Notes

### Competing Interest Statement

The authors have declared no competing interest.

### Funding Statement

Funding provided by Clemson University.

## REFERENCES

1. Dhama K, Khan S, Tiwari R, et al. Coronavirus Disease 2019–COVID-19. Clin Microbiol Rev 2020;33(4):e00028–20.

2. Hart OE, Halden RU. Computational analysis of SARS-CoV-2/COVID-19 surveillance by wastewater-based epidemiology locally and globally: Feasibility, economy, opportunities and challenges. Sci Total Environ 2020;730:138875.

3. Daughton CG. Wastewater surveillance for population-wide Covid-19: The present and future. Sci Total Environ 2020;736:139631.

4. Choi PM, Tscharke BJ, Donner E, et al. Wastewater-based epidemiology biomarkers: Past, present and future. TrAC - Trends Anal Chem 2018;105:453–69.

5. Chen C, Gao G, Xu Y, et al. SARS-CoV-2-Positive Sputum and Feces After Conversion of Pharyngeal Samples in Patients With COVID-19. Ann Intern Med 2020;172(12):832–4.

6. Chen Y, Chen L, Deng Q, et al. The presence of SARS-CoV-2 RNA in the feces of COVID-19 patients. J Med Virol 2020;92(7):833–40.

7. Wu F, Xiao A. SARS-CoV-2 titers in wastewater foreshadow dynamics and clinical presentation of new COVID-19 cases. J Chem Inf Model 2020;53(9):1689–99.

8. Kumar M, Patel AK, Shah A V., et al. First proof of the capability of wastewater surveillance for COVID-19 in India through detection of genetic material of SARS-CoV-2. Sci Total Environ 2020;746:141326.

9. Ahmed W, Angel N, Edson J, et al. First confirmed detection of SARS-CoV-2 in untreated wastewater in Australia: A proof of concept for the wastewater surveillance of COVID-19 in the community. Sci Total Environ 2020;

10. La Rosa G, Iaconelli M, Mancini P, et al. First detection of SARS-CoV-2 in untreated wastewaters in Italy. Sci Total Environ 2020;736:139652.

11. Sherchan SP, Shahin S, Ward LM, et al. First detection of SARS-CoV-2 RNA in wastewater in North America: A study in Louisiana, USA. Sci Total Environ 2020;743:140621.

12. Peccia J, Zulli A, Brackney DE, et al. SARS-CoV-2 RNA concentrations in primary municipal sewage sludge as a leading indicator of COVID-19 outbreak dynamics. medRxiv 2020.05.19.20105999; https://doi.org/10.1101/2020.05.19.20105999.

13. Godio A, Pace F, Vergnano A. Seir modeling of the italian epidemic of sars-cov-2 using computational swarm intelligence. Int J Environ Res Public Health 2020;17(10).

14. Carcione JM, Santos JE, Bagaini C, Ba J. A Simulation of a COVID-19 Epidemic Based on a Deterministic SEIR Model. Front Public Heal 2020;8(May).

15. He S, Peng Y, Sun K. SEIR modeling of the COVID-19 and its dynamics. Nonlinear Dyn 2020;0123456789.

16. Hethcote HW. Mathematics of infectious diseases. SIAM Rev 2000;42(4):599–653.

17. van den Driessche P. Reproduction numbers of infectious disease models. Infect Dis Model 2017;2(3):288–303.

18. Abou-Ismail A. Compartmental Models of the COVID-19 Pandemic for Physicians and Physician-Scientists. SN Compr Clin Med 2020;

19. Lauer SA, Grantz KH, Bi Q, et al. The incubation period of coronavirus disease 2019 (CoVID-19) from publicly reported confirmed cases: Estimation and application. Ann Intern Med 2020;172(9):577–82.

20. CDC. Duration of Isolation and Precautions for Adults with COVID-19. Centers Dis. Control Prev.2020;

21. Fang Y, Nie Y, Penny M. Transmission dynamics of the COVID-19 outbreak and effectiveness of government interventions: A data-driven analysis. J Med Virol 2020;92(6):645–59.

22. Liu Y, Gayle AA, Wilder-Smith A, Rocklöv J. The reproductive number of COVID-19 is higher compared to SARS coronavirus. J Travel Med 2020;27(2):1–4.

23. Wang Y, Wang Y, Chen Y, Qin Q. Unique epidemiological and clinical features of the emerging 2019 novel coronavirus pneumonia (COVID-19) implicate special control measures. J Med Virol 2020;92(6):568–76.

24. Wölfel R, Corman VM, Guggemos W, et al. Virological assessment of hospitalized patients with COVID-2019. Nature 2020;581(7809):465–9.

25. He X, Lau EHY, Wu P, et al. Temporal dynamics in viral shedding and transmissibility of COVID-19. Nat Med 2020;26(5):672–5.

26. Kapo KE, Paschka M, Vamshi R, Sebasky M, Mcdonough K. Estimation of U. S. sewer residence time distributions for national-scale risk assessment of down-the-drain chemicals. Sci Total Environ [Internet] 2017;603–604:445–52. Available from: https://doi.org/10.1016/j.scitotenv.2017.06.075

27. Hart OE, Halden RU. Modeling wastewater temperature and attenuation of sewage-borne biomarkers globally. Water Res 2020;172:115473.

28. Ahmed W, Bertsch PM, Bibby K, et al. Decay of SARS-CoV-2 and surrogate murine hepatitis virus RNA in untreated wastewater to inform application in wastewater-based epidemiology. Environ Res 2020;(August).

29. Rose C, Parker A, Jefferson B, Cartmell E. The characterization of feces and urine: A review of the literature to inform advanced treatment technology. Crit Rev Environ Sci Technol 2015;45(17):1827–79.

30. Wu SL, Mertens AN, Crider YS, et al. Substantial underestimation of SARS-CoV-2 infection in the United States. Nat Commun [Internet] 2020;11(1). Available from: http://dx.doi.org/10.1038/s41467-020-18272-4

