## Supplementary Appendix for "COVID-19 Wastewater Epidemiology: A Model to Estimate Infected Populations"

### SARS-CoV-2 DETECTION AND QUANTIFICATION

Sewage samples (225 mL) were prepared for analysis by pasteurization at 60 ºC for 30 minutes. The pasteurized samples were centrifuged at 6,500 g for 10 minutes at 6 ºC to remove solids. The supernatant was decanted into a chilled beaker in an ice bath. Reagent grade sodium chloride and polyethylene glycol 6000 (PEG) were added directly into the chilled supernatant with constant stirring until all the salt and PEG dissolved to achieve a final concentration of 2.3% salt w/v and 7% PEG. Virus was allowed to precipitate for several hours at 4 ºC. The virus was then collected in the pellet produced by centrifuging at 15,000g. The supernatant was decanted and the pellet was dissolved in 2 mL of Tris-EDTA-Salt (TES). The RNA in resuspended virus pellet was extracted in TRIzolTM reagent (ThermoFisher Scientific, Waltham, MA). The resulting RNA extract was prepared for RT-qPCR using a NucleoMag® Pathogen RNA/DNA Isolation Kit (Macherey-Nagel, Düren, Germany).

RNA was quantified by RT-qPCR using open reading frame 1ab (orf-1ab), nucleocapsid (N) protein, and spike (S) protein gene primers. MS2 bacteriophage RNA matrix spikes acted as an internal positive control for the RNA isolation and extraction procedures. MS2 phage RNA added to the RT-qPCR reaction acted as a PCR positive control. Additions of plasmids containing SARS-CoV-2 N gene fragments were used in low N gene sequence (1,000 copies per reaction) and high N gene sequence (10,000 copies per reaction) positive controls for the N gene primers. Purified SARS-CoV-2 RNA was used as low concentration (50 copies per reaction) and high concentration (250 copies per reaction) RNA positive controls. A no template negative control containing only nuclease free water controlled for contamination during the RNA extraction and RT-qPCR assembly. Quantification of gene copies/L was performed using the N protein gene. Detection limits ranged from 860 to 4,000 copies $L^{-1}$.

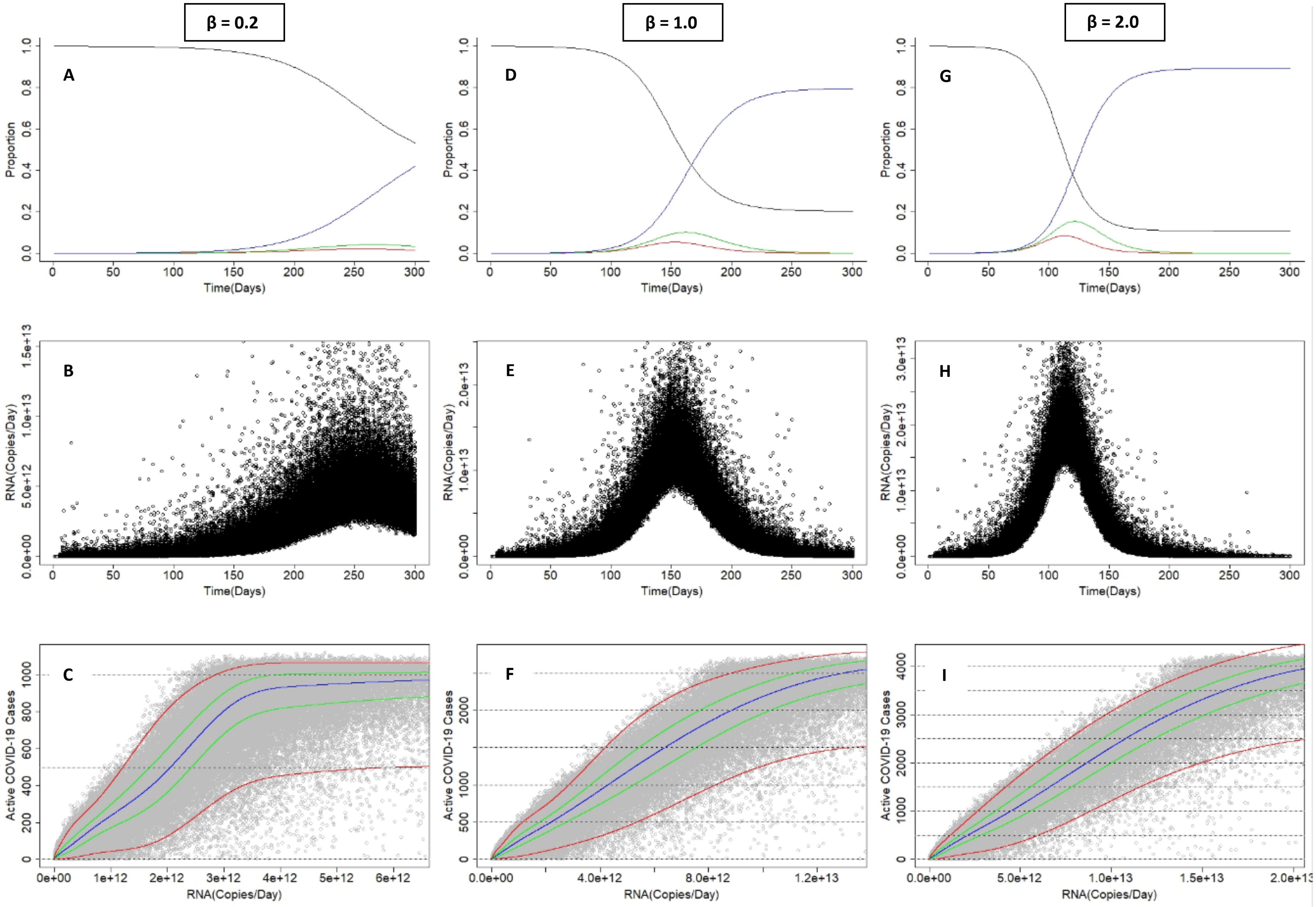


Figure S1: SEIR model for three β values. Panels A, D, G: Proportions of the population that are susceptible (**black**), exposed (**red**), infectious (**green**), and recovered (**blue**). Panels B, E, H: Model predictions for mass rate of SARS-CoV-2 RNA in wastewater over time. Individual black points represent each stimulation. Panel C, F, I: Predictions of the number of infections versus RNA mass rate. Individual gray points represent each stimulation. The **blue** line represents the median, the **green** and **red** lines represent the 75% and 95% confidence intervals, respectively.
